# Complementary frailty and mortality prediction models on older patients as a tool for assessing palliative care needs

**DOI:** 10.1101/2021.01.22.21249726

**Authors:** Vicent Blanes-Selva, Ascensión Doñate-Martínez, Gordon Linklater, Juan M. García-Gómez

**Affiliations:** Biomedical Data Science Lab, Instituto Universitarios de Tecnologías de la Información y Comunicaciones (ITACA), Universitat Politècnica de València, Valencia, Spain; Polibienestar Research Institute, University of Valencia, Spain; Highland Hospice and NHS Highland, Inverness, UK

**Keywords:** Palliative Care, Machine Learning, Deep learning, Frailty, Mortality, Admission, Needs assessment

## Abstract

**Introduction:** Palliative care (PC) has demonstrated benefits for life-limiting illnesses. Nowadays, there is a growing consensus about giving access these care services to non-cancer older patients. Bad survival prognosis and patients’ decline are working criterions to guide PC decision making.

**Objective:** The main aim of this work is to propose complementary models based on machine learning approaches to predict frailty and mortality in older patients in the context of supporting PC decision making.

**Methods:** The dataset used in this study is composed by 39,310 hospital admissions for 19,753 older patients (age >= 65) from January 1st, 2011 to December 30th, 2018. Predictive models based on Gradient Boosting Machines and Deep Neural Networks were implemented for binary one-year mortality classification, survival estimation and binary one-year frailty classification. Besides, we tested the similarity between mortality and frailty distributions.

**Results:** The one-year mortality classifier achieved an AUC ROC of 0.87 [0.86, 0.87]; whereas the mortality regression model achieved an MAE of 333.13 [323.10, 342.49] days. Moreover, the one-year frailty classifier obtained an AUC ROC of 0.89 [0.88, 0.90].

**Conclusions:** The performance of our one-year mortality model is competitive with the current state-of-the-art. Besides, to our knowledge, this is the first study predicting one-year frailty status based on a frailty index. We found mortality and frailty criteria are weakly correlated and have different distributions; therefore, we interpreted them as complementary assessment measurements for palliative care decision making. Predictive models are accessible as an online tool at http://demoiapc.upv.es. The models presented here may be part of decision support systems for care services in non-cancer older patients after their external validation.

## Introduction

Palliative Care (PC) is a holistic approach that improves the quality of life of patients with life-limiting diseases. It is recommended that it be incorporated early in the disease trajectory, even in conjunction with potentially curative treatments [1]. PC can improve quality of life [2], mood [3], symptom control [4], reduce emergency department visits and hospitalisation [5], and even increase the one-year survival [6].

PC services have traditionally been mainly accessed by cancer patients but there is growing consensus about the importance of promoting access for patients with non-malignant disease at earlier stages [7, 8, 9]. Patients’ prognosis and functional decline are two important elements in decision-making to be considered by health and care professionals in the introduction of PC needs assessment and/or PC conversations with older people.

On the one hand, it is estimated that at least 75% of patients would benefit from access to PC during their terminal illness [10]. Nevertheless, uncertainty about prognostication is cited as a common barrier to PC referral, particularly for patients with non-malignant disease [11]. On the other hand, frailty in older patients is defined as a state characterised by reduced physiological reserve and loss of resistance to stressors caused by accumulated age-related deficits [12]. Two of the most popular frailty dimensions are the frail phenotype by Fried et al. [13], which describes frailty as a biological syndrome; and the Frailty Index (FI) by Mitnitski et al. [14] which is based on health deficits accumulations, also, frailty has been defined since a more comprehensive approach taking into consideration a holistic understating of the person. In this sense, frailty can be experienced by a decrease in human functioning not only at physical level, but also at psychological and social domains [15]. Raudonis et al. [16] suggest in their study that frail older adults could benefit from involvement in PC programs as frailty is associated with poor health outcomes and death [17].

Different strategies have been used to try to aid prognostication. Clinical intuition was harnessed with the Surprise Question (“Would I be surprised if this patient died in the next year?”) which has been promoted as a tool to prompt clinicians to recognise patients with a limited prognosis [18] However, in 2017 Downar et al. [19] published a systematic review of the surprise question, concluding that more accurate tools are required given its poor to modest performance as a mortality predictor. Also, it has been demonstrated that the risk of death increases with lower performance levels and with falling performance levels, but survival data varied across different healthcare systems [20]. In this line, the Supportive and Palliative Care Indicators Tool (SPICT) proposes a set of clinical indicators of poor prognosis developed through a consensus of expert opinion [21], which has shown to have a predictive accuracy of up to 78% [22]. Other studies have used data analysis to propose alternative tools to predict short-term mortality. Bernabeu-Wittel in 2010 developed the PROFUND index [23], a predictive model for patients with multimorbidity. Van Walraven et al. in 2015 proposed HOMR [24], a tool for predicting one-year mortality on adults (>=18 years and >= 20 years for the different cohorts) In 2018 Avati et al. [25] proposed a deep learning approach to identify patients with a survival between 3 and 12 months a, in 2019 Wegier et al. [26] proposed a version of HOMR but using only variables available at the admission.

Additionally, and as stated before, quantifying frailty is important since as patients become frailer advance care planning conversations should be prioritized to establish patient goals and wishes in advancing serious illness [27], which may include the involvement in PC programmes. A wide array of frailty indexes has been proposed to assess the health status in older adults. Frailty index has been used as a tool to predict mortality and poor health outcomes [28]. Some studies have tried to predict frailty status: Babič et al. in 2019 [29] use a clustering approach to identify clusters considering the prefrail, non-frail and frail status using 10 numerical variables for adults over 60 years old. Sternberg et al. [30] in 2012 tried to identify frail patients with their methods against the VES frailty score [31] for patients over 65 years old. Bertini et al. [32] in 2018 created two predictive models for patients over 65 years old: one to assess frailty risk using the probability of hospitalization or death within the year and a second one to assess worsening risk to each subject in the lower risk class.

Based on these previous results, our aim in this work is to propose a machine learning tool capable of making predictions about mortality and frailty for older adults so health professionals can benefit from quantitative approaches based on data-driven evidence when deciding advance care planning. In this sense, we propose the creation of three different but complementary models: a) a one-year mortality classifier that will work as a binary predictor; b) a survival regression model aimed to obtain a prediction in days from admission to death; and c) a one-year frailty classifier to predict the health status, assessed by the Frailty Index, of a patient one year after admission. The authors consider that the combination of both mortality and frailty criteria, working as complementary information sources, can have a positive impact on detecting needs to start PC conversations.

## Materials

### Basic description

The dataset contained hospital admissions records for older patients (age >= 65) from January 1st, 2011 to December 31st, 2018. Patients admitted to psychiatry and obstetrics services were excluded from the study. Data was extracted from the system on November 1st, 2019.

Data contains a total of 39,310 hospitalization episodes corresponding to 19,753 unique patients. Cohort was composed by 9780 males and 9973 females with a mean age of 80.75 years (see Table 1).

**Table 1:** Patient demographic information.

| Sex | N-individuals | Mean Age (years) | STD Age (years) |
| --- | --- | --- | --- |
| Female | 9973 | 80.75 | 8.67 |
| Male | 9780 | 77.44 | 8.24 |
| All | 19753 | 79.11 | 8.62 |

### Mortality target variables

Mortality target variables were extracted from admission administrative data and recorded death date. Patients without a record of death were considered alive at the time of extraction and consequently were excluded from the one-year mortality model development unless a whole year passed since the admission date. Those patients alive at the extraction date were not eligible for the survival regression problem, which is known as right-censored data.

### Frailty target variable

As for the frailty target, following the work of Searle et al. [33] we calculated the frailty index (FI) of every episode (admission frailty) and sorted them chronologically from oldest to newest. The target FI was the admission frailty of the following episode in chronological order if this next episode happened within the year. If a patient had multiple admissions during the following year, we used the most recent episode as target. Otherwise, target frailty was set to the same value as the current admission frailty. Most recent episodes and patients with only one episode were removed because no posterior data was available, so we considered them as censored data.

Finally, we stratified the FI in 4 categories according to the work of Hoover et al. [34] and aggregate together the two less severe frailty conditions (Non-Frail + Vulnerable) and the two more frail status (Frail + Most Frail). Variables used in the frailty index are listed in Table 2 and were extracted as part of the original 147 variables.

**Table 2:** List of variables included in the frailty index and their distribution. All variables are binary, and their distribution represents the presence (Y) or absence (N) of the condition.

| **Variable** | **Distribution** | **Variable** | **Distribution** |
| --- | --- | --- | --- |
| Difficulties in dressing | Y: 3829, N: 35481 | Difficulties in urinating | Y: 3683, N: 35627 |
| Difficulties in bathing | Y: 5999, N: 33311 | Difficulties in stooling | Y: 3121, N: 36189 |
| Difficulties in grooming | Y: 5242, N: 34068 | Difficulties in eating | Y: 2965, N: 36345 |
| Difficulties in moving | Y: 3398, N: 35912 | Hypertension | Y: 30975, N: 8335 |
| COPD | Y: 9724, N: 29586 | Heart Failure | Y: 13228, N: 26082 |
| Stroke | Y: 9828, N: 29482 | Parkinson | Y: 1655, N: 37655 |
| Thyroid Disorders | Y: 4538, N: 34772 | Diabetes Mellitus | Y: 15910, N: 23400 |
| Gastrointestinal or liver disease | Y: 27401, N: 11909 | Musculoskeletal Diseases | Y: 24330, N: 14980 |
| Dementia | Y: 4479, N: 34831 | Malnutrition | Y: 2718, N: 36592 |
| Pressure Ulcers | Y: 1886, N: 37424 | Anaemia | Y: 12546, N: 26764 |
| Hear impairment | Y: 6777, N: 32533 | Gastrointestinal problems | Y: 12567, N: 26743 |
| Chronic renal failure | Y: 8679, N: 30631 | Depression | Y: 587, N: 38723 |
| Cancer | Y: 16536, N: 22774 | Constipation | Y: 5088, N: 34222 |
| Atrial fibrillation | Y: 12434, N: 26876 | Visual impairment | Y: 20100, N: 19210 |
| Psychiatric disease | Y: 19436, N: 19874 |  |  |

### Data censoring and distributions

After data censoring, the one-year mortality target variable distribution was: 24985 (65.83%) episodes were negative cases (time to exitus > 365 days), and 13431 (34.17%) episodes were positive (time to exitus <= 365 days) as shown in Figure 1.A. The survival regression target variable (20959 episodes; mean 368.59; range [0 to 3033]) presents a right-skewed shape as can be observed in its density plot in Figure 1.B.

**Figure 1:**
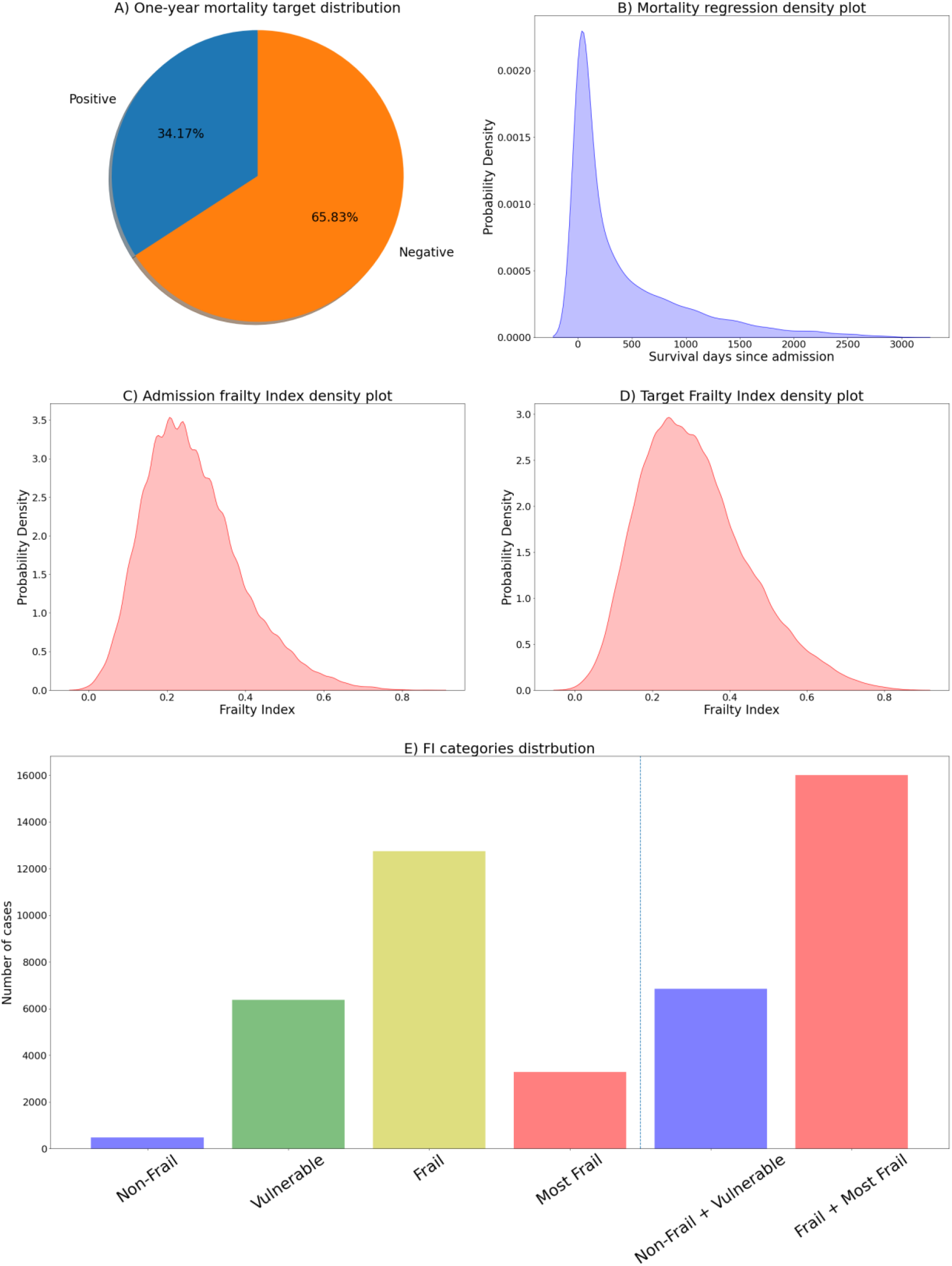
A) One-year mortality target distribution; B) Density plot from survival regression target variable; C) Density plot from the FI target variable; D) Density plot from the admission FI; E) FI categories distribution.

The admission FI (mean 0.27; std 0.12) and the FI target variable (22859 episodes; mean 0.32; std 0.14), resembled a slightly skewed normal distribution (plot in Figure 1.C and Figure 1.D), while the distributions of the different categories are: Non-Frail 986 (2.2%), Vulnerable 10911 (24.34%), Frail 25638 (57.19%), Most Frail 7294 (16.27%). As aggrupation of two categories: Non-Frail + Vulnerable 11897 (26.54%), Frail + Most Frail 32932 (73.46%), data represented in Figure 1.E.

## Methods

### Predictive models

As first approach for predictive models, we have selected the Gradient Boosting Machines (GBM) [35], which can be used for both classification and regression. GBMs are ensemble models composed by decision trees. This model follows a training iterative algorithm, where in each step the tree that minimizes the selected loss function is added to the ensemble until the hyperparameter setting the number of trees is reached. The GBM models are known for their notable performance on different problems [36, 37, 38].

Our second approximation to the predictive models is through the Deep Neural Network (DNN) [39]. Due to the tabular nature of the data, we are using a multilayer perceptron topology, which is composed by interconnected neurons. The neurons are connected by weights and their output is a function of the sum of the inputs to the neuron, applying a non-linear activation function afterwards [40]. Our models are using Batch Normalization [41] and Dropout [42] as regularization method and the Leaky ReLU [43] function as activation function. Deep learning has been a trendy technology when dealing with the increasing amount of data, and its application to medicine is growing [44].

### Hyperparameters and variable selection

To select the hyperparameters and make the selection of variables, we split the datasets (80%/20%) into a design set and an evaluation set. Then, we used a recursive feature elimination process as a filter method on the design set. This process starts with the whole set of features, trains a tree-based model, calculates the relevance of each variable using the GINI importance [45], which measures the average gain of purity in the tree splits. Finally, less relevant variables are eliminated. The process is repeated until the desired number of features is obtained. The number of variables was set to 20 in each task, a number of variables able to be handled by a human operator, with 2 variables eliminated each iteration. Table 8 on supplementary material provides a description for each selected variable.

The selection of hyperparameters for each model was performed using the Optuna optimization library [46]. Using this approach, we selected the most relevant hyperparameters for the GBM and the DNN and provide feasible ranges. During the process, the method selects a value for each hyperparameter, train the model with the 80% of the design set and evaluate it with the remaining 20% and the appropriate metric. As more iterations occur, Optuna make a smarter selection of the hyperparameters until the algorithm reach a selected number of iterations.

### Evaluation

To evaluate the models, we used the bootstrap technique [47] with 1000 resamples on the unseen evaluation set. To evaluate the performance of the one-year mortality and the frailty binary classifier we selected the following metrics: area under ROC curve (AUC ROC), accuracy, sensitivity (or True Positive Rate) and specificity (or True Negative Rate). For the survival regression model, we selected the mean absolute error (MAE), in addition, we repeated the regression experiments using only those cases where the prediction is < 500 days. In addition, since the GBM is an explicable model, we reported the contribution of each variable in percentage.

### Comparison with baseline models

To compare our mortality regression model with state of the art, we have performed survival analysis over the data processed with same pipelines described above. For that, we chose the Cox regression model [48], from which we obtained survival estimations for patients by calculating the survival expected time. To compare the classification models, we trained a binary Logistic Regression for both mortality and frailty.

### Software

The whole experimentation described in this work has been carried using the python 3 programming language [49], and the following scientific libraries and packages: numpy as the main mathematical library [50], pandas’ data frames to handle the data representation [51], scikit-learn’s implementation of GBM [52], Pytorch’s DNN implementation [53], Optuna as hyperparameters selection [46] and lifelines’ implementation of the Cox model [54].

## Results

### Associations between distributions

The Spearman’s correlation coefficient between the survival target in days and the admission FI was -0.10 while the correlation between survival and the target FI was -0.16, both correlations were statistically significant (p < 0.001). The similarity between the binary 1-year mortality target and the binary FI target was studied using the Fisher’s exact test, rejecting the null hypothesis (p < 0.001), and therefore concluding both distributions lack non-random associations.

### One-year morality classifier

GBM and DNN performed very closely (0.87 CI 95% [0.86, 0.87] and 0.86 CI 95% [0.85, 0.86] AUC ROC), both outperforming the logistic regression baseline, complete results and metrics on Table 4.

**Table 4:**
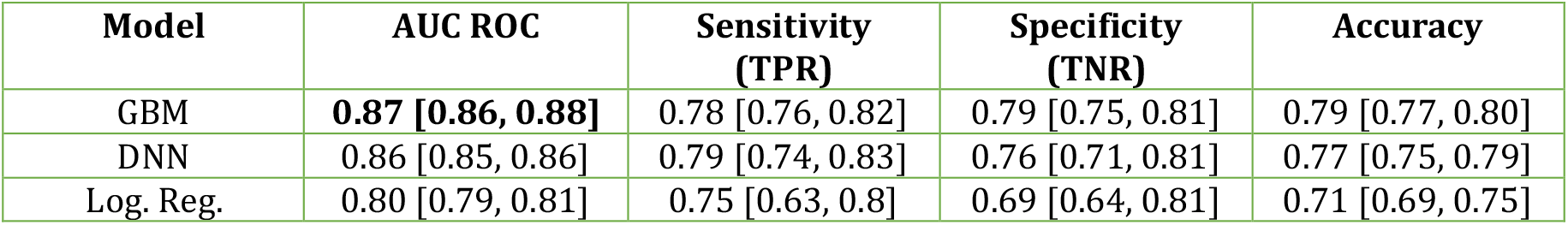
One-year mortality classifier evaluation. Reporting the mean and the 95% confidence interval

### Survival regression

The cox regression produced a MAE of 444.8 days while the GBM and the DNN model achieved a MAE of 333.13 and 338.88 days respectively. The GBM outperformed the rest of the models when using only samples with survival < 500, complete performance for survival regression models on Table 5.

**Table 5:** Mortality regressor evaluation. Reporting the mean and the 95% confidence interval.

| Model | MAE | MAE (<500d) |
| --- | --- | --- |
| GBM | **333.13 [323.10, 342.49]** | **94.67 [92.02, 97.49]** |
| DNN | 338.88 [329.07, 349.37] | 103.21 [100.47, 106.08] |
| Cox | 444.8 [438.9, 450.9] | 116.71 [115.23, 118.08] |

### One-year frailty classifier

The classification model based on the logistic regression achieved an AUC ROC of 0.84, while the GBM and DNN outperformed it with an AUC ROC of 0.89. Complete metrics for the frailty classification are available in Table 6.

**Table 6:** Frailty classifier evaluation

| Model | AUC ROC | Sensitivity (TPR) | Specificity (TNR) | Accuracy |
| --- | --- | --- | --- | --- |
| GBM | **0.89 [0.88, 0.90]** | 0.77 [0.73, 0.81] | 0.85 [0.81, 0.89] | 0.79 [0.78, 0.81] |
| DNN | **0.89 [0.88, 0.90]** | 0.76 [0.72, 0.83] | 0.85 [0.78, 0.89] | 0.79 [0.77, 0.82] |
| Logistic Reg. | 0.84 [0.83, 0.85] | 0.74 [0.70, 0.78] | 0.78 [0.73, 0.83] | 0.75 [0.73, 0.77] |

### GINI Importances

Following the previous methodology, we have calculated the GINI importance for each of the GBM predictive models. For the one-year mortality model, the most important variables were: Number of Active Groups, Charlson Index and Age. In the regression task: Number of Active Groups, Charlson Index and Service whereas in the model version including only cases with survival < 500 days were: Leukocytes, C-reactive protein and Urea. Finally, the most relevant features in the frailty model were the Charlson Index, Number of previous Emergency Room visits and Hypertension. Complete details in Table 7.

**Table 7:**
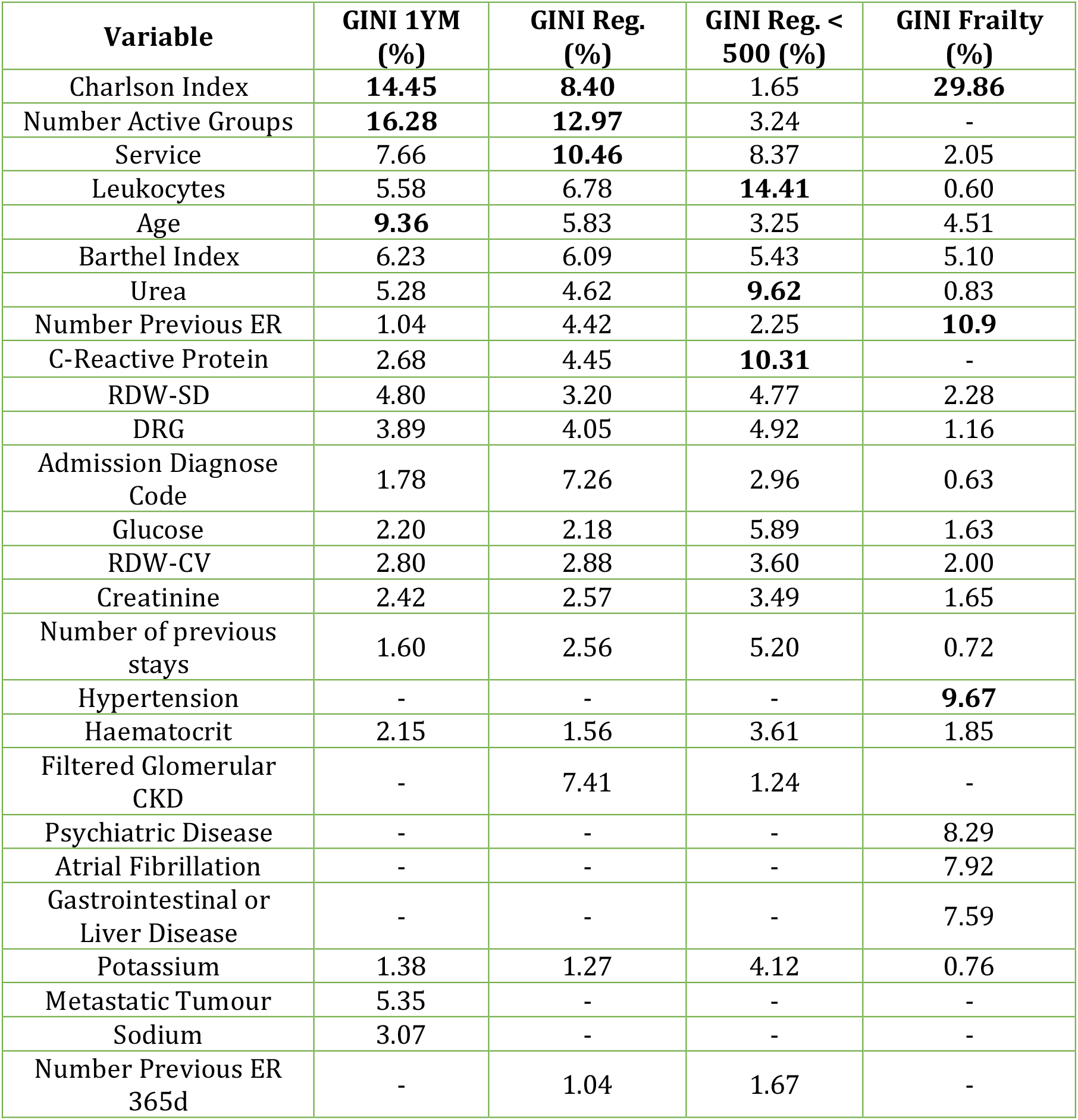
GINI importance of the GBM for mortality and frailty tasks. Variables are sorted using the sum of the GINI importances in all tasks.

## Discussion

The overall aim of this study was to develop machine learning tools capable of making predictions about mortality and frailty for older adults so that health professionals can benefit from quantitative approaches based on data-driven evidence. First, we created a one-year mortality model working as a binary predictor. Additionally, a survival regression model was created aimed to obtain a prediction in days from admission to death. Finally, a frailty model to make predictions for 12 months after the admission.

Our one-year mortality model ranked among the best general admission models in terms of AUC ROC (0.87 CI 95% [0.86, 0.88]). Outperforming PROFUND (0.77) [23] and scoring slightly below HOMR (0.89-0.92) [24], mHOMR (0.89) [26] and Avati’s deep learning approach (0.93, 0.87 for admitted only patients) [Avati18]. However, our model is not fully comparable since it targeted older adults (>= 65 years old) meanwhile all the mentioned studies use inclusion criteria of >= 18, except Avati, which also includes paediatric records. Yourman et al. [55] reviewed prognosis indices for older patients, where the better AUC ROC for 1-year index was 0.83, which is below that out lower 95% CI bound. As expected, the GBM model performed significantly better than the Logistic Regression counterpart and slightly better than the DNN model.

Our survival regression model scored a mean absolute error of 329.97 days, outperforming the 444.8 days scored by the cox model. Despite obtaining better predictions than one of the most used models when dealing with survival time, a mean error of almost a year does not seem to adequately meet the original purpose of this model. When removing cases where survival time is greater than 500 days, the GBM performs better than the other models achieving a mean error of 94.67 days; this improves the prediction error and will be likely better accepted by the health care professionals. We have selected this later model based on GBM to be included in the online tool. This improvement in the predictive power is likely due to removing the long tail in the distribution that includes infrequent values and outliers. It would also be possible to train a model using cases where survival was less than 365 days. In this case, the model would be used only when the one-year mortality produces a positive result; a preliminary result using the GBM configuration produced an MAE of 69.89 CI 95% [67.83, 72.08]. A further study concerning health care experts’ preferences is needed to know if this alternative is preferred over the standard approximation.

The 1-year frailty model scored a 0.89 AUC ROC on both GBM and DNN, outperforming the logistic regression version (0.84 AUC ROC). These results demonstrate a great predictive power for assessing a patient’s frailty index category one year from admission. As far as authors know, this is the first study where a model is used to predict a future frailty status without using proxies such as mortality or disability. These models use variables containing information about the current frailty status combined with other factors such as the previous stays in the emergency room or the age to determine the future frailty status. Since most of the variables are shared with the other two mortality models, the addition of few extra variables means that we can predict the future frailty status with a low additional effort.

Each model was set with the 20 most relevant variables from a total of 147, a number that was arguably too high to be used by a human operator. This selection was performed using the Random Forest’s GINI importance criteria with recursive feature elimination as a data-driven method. This method is known to have favour bias towards categorical variables with large number of categories and continuous variables, however it is widely used because it is simple and fast to compute [45]. In the end, all three models share a great number of variables (Table 7), being only 26 different variables. The selected variables by the recursive feature elimination algorithm are coherent with the different mortality works in the literature [23, 24].

These results provide a complementary perspective based on an objective measure of frailty to initiate early PC. The mean admission FI was 0.27 ± 0.12, and its shape resembles a normal distribution. This is a coherent behaviour with the findings in the Mitnitski et al. study [14], where the most impaired groups have a bigger FI mean, and the distribution is shaped like a normal distribution, as opposed to the less impaired groups which had a smaller mean FI and can be approximated using a gamma distribution. The correlation between our admission FI and MR target in days is –0.10, lower than the one reported in [14] which was –0.234. These means that the frailty index used in this work, for this sample, is less associated with mortality.

The relationship between frailty and mortality have been studied previously [28] pointing to the association between both. Despite the similarity in the used input variables, the target variable distributions are poorly correlated and have different shapes. Both criteria have been highlighted as important for accessing PC in previous studies and are related. However, they reflect two different distributions, and the authors think of them as two complementary criteria. Therefore, we conclude that the best approximation for taking advantage of both mortality and frailty criteria is to have different predictive models working at the same time, increasing the information to support the decision-making process. The incorporation of the criterion of frailty may represent an added value for those health professionals deciding about inclusion in PC services. This is in line with results obtained by Almagro et al. (2017) [56] showing that the use of poor vital prognosis as exclusive criteria for initiating PC among COPD patients should be critically appraised.

This study’s clinical impact resides in the potential to predict negative outcomes for hospital admitted patients within the next year. First, we choose one year as a horizon to make the mortality prediction, as stated elsewhere [25], longer than 12 months is not desirable due to the difficulty in the predictions and the limited resources of the programs which are better to focus on immediate needs. Thus, referral to PC may be focused on immediate needs. Also, despite being more difficult to predict, the information provided by the survival regression model may help to contextualise the results of the one-year mortality model. Therefore, health professional would be supported with additional information such as the magnitude of the remaining time until death in terms of days, weeks or months. The inclusion of these models into clinical practise could help anticipate the decline in admitted patients, allowing health professionals to allocate scarce resources to patients that will need them the most.

The main contribution of this work is the development of the mortality and frailty models. We have constructed machine learning mortality models that can predict accurately the death of older admitted patients. In addition to this, we proposed a predictive model to assess the future frailty status of the patients. All the three models were implemented in an online tool [57] that is available to any health care expert for academic use until further validations. In this work we demonstrate the complementariness of the mortality and frailty models testing the null correlation between both factors in our dataset so we should treat them as complementary criterions.

The main limitation of this study is the use of data from only one hospital. Therefore, internal validation only assures the performance of the models with similar data. We cannot ensure the reported efficiency in other hospitals and/or with other patient populations [58]. Also, data from the same centres can change over time due a wide variety of reasons such as change in protocols or external agents such a pandemic [59, 60]. Additional external validations are needed for future work. Broader populations can be approached implementing predictive models using EHR, supporting an effective identification of patients in need of further specialized care [61]. Thus, besides external validation of the models, future authors’ work will require a significant software development and implementation project to connect these systems with hospital EHR and avoid manual input by professionals. Also, the maturity of the models and the software wrapping them needs to be *field tested* before their inclusion as standard tool to the hospital information system.

## Conclusion

This work proposes the use of three different machine learning models based on hospital admission data to assess PC needs on older adults and help healthcare professionals in the decision-making process. The authors constructed three different but complementary predictive systems: a one-year mortality model, a regression mortality model to provide more information about the first prediction, and a one-year frailty model. There are previous modern mortality models using machine learning methods available elsewhere, but they are not specifically focused on older populations. Also, to our knowledge, this is the first study predicting one-year frailty status based on a frailty index. As previous studies have shown, mortality and frailty could be relevant criteria to admit patients to PC programs.

Therefore, health professionals could benefit from the use of data-driven accurate predictions of these two dimensions on patients over 65. In addition to the benefits experienced by patients and their families, the early identification of these patients’ needs can help better manage the available health and social care resources and may even reduce costs overall. Consequently, the authors propose the use of predictions in both mortality and frailty as complementary predictions to help assess PC needs due to its individual relevance but weak correlation, reliability and great predictive power. The described models have been implemented and publicly available for academic purpose at [57]

### Ethics

The data used in this study comes from the University and Polytechnic La Fe Hospital of Valencia and was retrospectively collected from the Electronic Health Records (EHR) of the hospital. This procedure was assessed and approved by the Ethical Committee of the University and Polytechnic La Fe Hospital of Valencia (registration number: 2019-88-1). Required patient patient-informed consent was waived. All methods were performed in accordance with the relevant guidelines and regulations.

## Supporting information

Table 8 on supplementary material provides a description for each selected variable

## Data Availability

The data is NOT publicly available

## Acknowledgements

The authors thank their contributions to María Soledad Giménez-Campos, María Eugenia Gas-López, María José Caballero Mateos and Bernardo Valdivieso. Special thanks to Ángel Sánchez-García for his contributions to the website.

## Conflicts of Interest

The authors declare that they have no competing interests.

## Abbreviations

PC: Palliative Care
FI: Frailty Index
ROC: Receiver Operating Characteristic
AUC ROC: Area Under the Curve ROC
GBM: Gradient Boosting Machine
1YM: One-year Mortality

## Notes

### Competing Interest Statement

The authors have declared no competing interest.

### Funding Statement

This work was supported by the InAdvance project (H2020-SC1-BHC-2018-2020 No. 825750.) and the ALBATROSS project (PID2019-104978RB-I00). Special thanks to Angel Sanchez-Garcia for his contributions to the website

### Author Declarations

This procedure was assessed and approved by the Ethical Committee of the University and Polytechnic La Fe Hospital of Valencia (registration number: 2019-88-1).

### Summary of Updates

Methodology reviewed to use bootstrap evaluation and added neural networks to compare the results of the original models

