## Supplementary material for "Complementary frailty and mortality prediction models on older patients as a tool for assessing palliative care needs": Table 8 on supplementary material provides a description for each selected variable

Table 8: Variables used in the predictive models and their descriptions.

| **Variable** | **Description** |
| --- | --- |
| Admission Diagnose Code | ICD9 code representing the main reason for the admission |
| Age | Patient's age |
| Atrial Fibrillation | ICD9 Diagnosis code: Atrial fibrillation (no/yes) |
| Barthel index | Barthel Index is an ordinal scale used to measure performance in activities of daily living (ADL). Ten variables describing ADL and mobility are scored, a higher number being a reflection of greater ability to function independently following hospital discharge. |
| Charlson index | The Charlson comorbidity index predicts the one-year mortality for a patient who may have a range of comorbid conditions, such as heart disease, AIDS, or cancer (a total of 17 conditions: Acute myocardial infarction, Congestive heart failure, Peripheral vascular disease, Cerebrovascular disease, Dementia, Chronic lung disease, Mild liver disease, Mild to moderate diabetes, Diabetes with chronic complications, Hemiparaplegia or paraplegia, Kidney disease, Malignant tumours, Moderate to serious liver disease, solid, metastatic tumour and AIDS). Each condition is assigned a score of 1, 2, 3, or 6, depending on the risk of dying associated with each one. |
| Creatinine | Lab result expressed in mg/dL |
| DRG | Diagnosis-related group (DRG) is a system to classify hospital cases into one of originally 467 groups |
| Filtered Glomerular CKD | Filtered Glomerular CKD lab result in ml/min/1,73 m² |
| Gastrointestinal or Liver Disease | ICD9 Diagnosis code: Gastrointestinal or Liver Disease (no/yes) |
| Glucose | Lab result expressed in mg/dL |
| Haematocrit | Lab result expressed in % |
| Hypertension | ICD9 Diagnosis code: Hypertension (no/yes) |
| Leukocyte | 10³/microL |
| Number Active groups | Number of active groups (medications) in each episode |
| Number of previous stays | Number of previous hospital admissions |
| Number Previous ER 365d | Number of previous Emergency Room visits (last 365 days) |
| Number Previous ER | Number of previous Emergency Room visits |
| Metastatic Tumour | ICD9 Diagnosis code: Metastatic tumour (no/yes) |
| PCR | C-Reactive protein lab result expressed in mg/L |
| Potassium | Lab result expressed in mEq/L |
| Psychiatric Disease | ICD9 Diagnosis code: Psychiatric disease (No/yes) |
| RDW-CV | The red cell distribution width (RDW) blood test measures the amount of red blood cell variation in volume and size. This values is the coefficient of variation of RDW |
| RDW-SD | Standard deviation of RDW measure |
| Service | Last Service updated during the stay |
| Sodium | Lab result expressed in mEq/L |
| Urea | Lab result expressed in mg/dL |
